# Sex-, age-, and BMI-specific patterns of visual and numerical body-size ideals in Japan: a large cross-sectional web-based survey

**DOI:** 10.64898/2026.10.19.26365487

**Authors:** Hiroki Tabata, Shota Sakamoto, Ko Fukumura, Mayuka Kanda, Yusuke Adachi, Yasuyo Yoshizawa, Yoshifumi Tamura

**Affiliations:** Sportology Center, Juntendo University, Graduate School of Medicine, 2-1-1 Hongo, Bunkyo-ku, Tokyo 113-8421, Japan; Juntendo Advanced Research Institute for Health Science, 2-1-1 Hongo, Bunkyo-ku, Tokyo 113-8421, Japan; Department of Sports Medicine & Sportology, Juntendo University, Graduate School of Medicine, 2-1-1 Hongo, Bunkyo-ku, Tokyo 113-8421, Japan; Research Fellow of the Japan Society for the Promotion of Science, 5-3-1 Kojimachi, Chiyoda-ku, Tokyo 102-0083, Japan; Research Institute for Bioscience Products & Fine Chemicals, Ajinomoto Co., Inc., 7-1 Kyobashi 1-Chome, Chuo-ku, Tokyo 104-0031, Japan; Faculty of International Liberal Arts, Juntendo University, 2-1-1 Hongo, Bunkyo-ku, Tokyo 113-8421, Japan; Department of Metabolism & Endocrinology, Juntendo University, Graduate School of Medicine, 2-1-1 Hongo, Bunkyo-ku, Tokyo 113-8421, Japan

**Keywords:** body image, thinness-oriented ideal, ideal BMI, silhouette, BMI status, age, sex, Japan

## Abstract

**Background:** Thinness-oriented body ideals are well described among young Japanese women, but variation in visual and numerical ideals by sex, age, and current BMI remains unclear.

**Methods:** We analysed an unweighted web-panel sample of 27,359 Japanese adolescents and adults aged 15–69 years from the Survey of Body Shape and Health Awareness in Japanese People, a cross-sectional web-based survey. Current and ideal BMI were derived from self-reported height and weight, and visual orientation from ideal versus perceived current silhouette. Adult BMI-stratified analyses included participants aged 20-69 years.

**Findings:** Among 27,359 participants (10,094 men and 17,265 women), relative thinness-oriented indicators increased with current BMI in both sexes. Among normal-weight women, 74·7-83·7% selected a smaller ideal silhouette and 79·2-91·2% reported an ideal BMI lower than current BMI. These indicators were also present among underweight women aged 20-29 years (44·9% and 43·4%, respectively), but declined with age. Most underweight men reported weight-gain-oriented ideals. Fine stratification showed that thinness-oriented ideals among women were common in low-normal and mid-normal BMI strata.

**Interpretation:** Body-size ideals differed by sex, age, and current BMI. Thinness-oriented ideals among women extended beyond obesity and young adulthood, whereas underweight men generally preferred a larger body. Longitudinal and behavioural studies are needed.

**Funding:** Cabinet Office of Japan Strategic Innovation Promotion Program (SIP), "Inclusive Community Platform" (grant no. SIPCP23C02).

**Research in context:** *Evidence before this study:* We searched Web of Science from database inception to May 11, 2026, when the search was updated for submission, using combinations of the terms “body image”, “body dissatisfaction”, “thinness”, “underweight”, “age”, and “sex”. We primarily considered articles published in English or Japanese. We did not restrict eligibility by study design or participant age or sex, but excluded studies conducted exclusively in patients with eating disorders, anorexia nervosa, or other psychiatric disorders. Previous studies from Japan mainly focused on adolescent girls and young women, whereas evidence covering men, middle-aged and older adults, and detailed current-BMI strata was limited. Most of the available evidence was cross-sectional, and heterogeneity in study populations, sampling methods, and measures of body image limited direct comparisons across studies.

*Added value of this study:* This large web-based survey maps visual and numerical body-size ideals simultaneously by sex, age, and current BMI in Japan. Fine BMI stratification shows that thinness-oriented ideals among women are already common in low-normal and mid-normal BMI strata, and the analysis highlights contrasting ideals among underweight women and men.

*Implications of all the available evidence:* Body-image-sensitive public-health research should extend beyond young women and should consider sex, age, and current BMI jointly. Longitudinal and intervention studies are needed to determine whether these ideals influence weight-control behaviour, nutritional status, body composition, or movement into and out of underweight.

## Introduction

Body size is one of the most visible physical characteristics of the human body and is associated with a wide range of health outcomes. Obesity is linked to type 2 diabetes, coronary heart disease, and a broad range of cardiometabolic comorbidities,^1^ whereas low body weight and undernutrition are associated with low bone mass, frailty, sarcopenia-related problems, and other adverse health outcomes across the life course.^2-4^ However, health outcomes may be associated not only with actual body size but also with body image. In particular, body dissatisfaction and discrepancies between actual, perceived, and ideal body size have been associated with dieting, disordered eating, depressive symptoms, and other health-related behaviours across adolescence and adulthood.^5-8^

These issues are especially relevant in Japan, where concerns related to low body weight are present at both ends of adulthood. Among young women, Japan has retained a distinctive profile of female underweight relative to many other countries. Based on the NCD-RisC age-standardised country estimates for 2022, Japan had the highest point estimate for the prevalence of underweight among adult women within the OECD (16·1%), followed by South Korea (7·9%).^9^ Domestically, the 2023 National Health and Nutrition Survey reported BMI <18·5 kg/m² in 20·2% of women in their 20s and 30s.^10^ Underweight in young women has been associated with reduced bone mass, menstrual dysfunction, and low-birthweight offspring, and has recently been reframed as a multi-system condition under the concept of Female Underweight/Undernutrition Syndrome (FUS).^11^ At the other end of adulthood, Japan is one of the world’s most aged societies, with 29·3% of the population aged ≥65 years in 2024.^12^ Low body weight and undernutrition in later adulthood are associated with frailty, sarcopenia, malnutrition, and increased mortality.^2,4,13,14^ Reflecting these dual challenges, Health Japan 21 (third term), Japan’s national health-promotion strategy, sets life-course targets to reduce underweight among young women and undernutrition among older adults.^15^

Previous Japanese studies have shown that thinness-oriented ideals are common among adolescent girls and young adult women, even among those whose current BMI is within or below the normal range.^16,17^ Consistent with these findings, Japanese high school girls have been shown to idealize body shapes substantially thinner than their current body shape.^18^ More broadly, body image may vary across the life course. In Western populations, age-related differences in body evaluation and body ideals have been described across adulthood,^5,7^ alongside age-related changes in body shape.^19^ However, corresponding evidence across a broad adult age range in Japan remains limited, and it is unclear whether patterns observed in Western populations apply directly to Japan.

Three aspects of the existing literature remain limited and motivate the descriptive design of the present study. First, the distribution of body-size ideals in midlife and early older adulthood has rarely been described, despite evidence that a persistent preference for thinness in later life may conflict with public-health messages emphasizing adequate nutrition and muscle maintenance to prevent sarcopenia and frailty.^2,4,5,7,13,20^ A description spanning adolescence to early older adulthood within a single sample is therefore needed. Second, body-size ideals in men have rarely been examined alongside those in women in comparable Japanese samples, although body-image dissatisfaction and appearance-related pressures are also reported among men.^21,22^ Concurrent description in both sexes is needed to identify where patterns converge or diverge. Third, body-size ideals have typically been measured either visually (silhouette tasks) or numerically (target weight or BMI), and their parallel distributions across sex, age, and current-BMI strata remain insufficiently described. In particular, whether thinness-oriented ideals are confined to participants with higher current BMI or are also common within the normal-weight range—and where along the BMI distribution the orientation begins to shift—has rarely been examined at a resolution that distinguishes low-normal from mid-normal BMI.

The present study therefore aimed to describe sex-, age-, and current-BMI-specific patterns of visual and numerical body-size ideals in a large Japanese web-based survey of adolescents and adults aged 15-69 years. To address the three gaps above, we first summarized visual and numerical body-size ideals by sex, age group, and conventional BMI status (underweight, normal weight, and obesity), with particular attention to normal-weight women, underweight women, and underweight men. We then characterized the within-category gradient of relative thinness-and weight-gain-oriented ideals across finer current-BMI strata. Together, these analyses were intended to provide a high-resolution descriptive map of body-size ideals across sex, age, and BMI strata in Japan.

## Methods

### Study design and setting

This study used data from the web-based component of the Survey of Body Shape and Health Awareness in Japanese People. Data collection was conducted between March 11 and March 25, 2024, through Rakuten Insight, Inc., which administered the survey to registered panel members. The deidentified analytic file contained completed response records only. Duplicate submissions from the same individual were prevented by vendor-assigned unique monitor IDs. Vendor-side invitation counts, view rates, participation rates, and page-by-page completion information were not available in the analytic file; available and unavailable web-survey reporting items are summarized in Supplementary Table S2. This report was prepared in accordance with the STROBE statement for observational studies and with reference to CHERRIES recommendations where applicable.^23,24^

### Participants and sampling strategy

Participants were Japanese adolescents and adults aged 15-69 years. Individuals who were unable to read and answer the questionnaire independently, minors younger than 18 years without proxy consent, individuals whose responses could not be accommodated by the binary sex-specific questionnaire branches used in the parent survey, and individuals with rare diseases affecting fewer than 50,000 patients in Japan were not eligible. Sample allocation was based on the Population Estimates of Japan as of October 1, 2022.²⁵ The estimates were used to approximate the demographic structure of the Japanese population through stratified web-panel allocation. In the first stage, 20,000 respondents were allocated across 564 strata defined by prefecture, sex, and age group. In the second stage, an additional 7,359 women aged 20-59 years were recruited using the same prefecture-by-age-group allocation framework to increase precision for analyses among women, yielding a combined sample of 27,359 respondents. The geographic distribution of participants across the 47 prefectures is shown in Supplementary Figure S1. This design supports sex-, age-, and BMI-specific description in a demographically allocated sample, particularly among women, but does not remove the limitations inherent to web-panel recruitment. Analyses were conducted without weighting; pooled percentages should therefore be interpreted as observations from a combined web-panel sample rather than nationally representative prevalence estimates. The target sample size was determined by the prespecified two-stage survey allocation; no formal sample-size calculation was performed.

### Body-size and body-image measures

Height, current weight, and ideal weight were self-reported. Current BMI and ideal BMI were calculated as weight in kilograms divided by height in meters squared. Numerical body-size orientation was categorized as ideal BMI lower than, equal to, or higher than current BMI; this directional classification captured only the direction, not the magnitude or motivation, of desired BMI change; ideal BMI <18·5 kg/m2 was also described as an absolute low-BMI ideal. For the BMI-status-specific analysis (Table 2), current BMI categories were defined as underweight (<18·5 kg/m2), normal weight (18·5-24·9 kg/m2), and obesity (≥25·0 kg/m2), using conventional adult underweight criteria and the Japanese obesity threshold.^26^ For the finer stratification analysis (Figure 1), current BMI was further divided into <18·5, 18·5-19·9, 20·0-22·4, 22·5-24·9, and ≥25·0 kg/m2.

**Figure 1.**
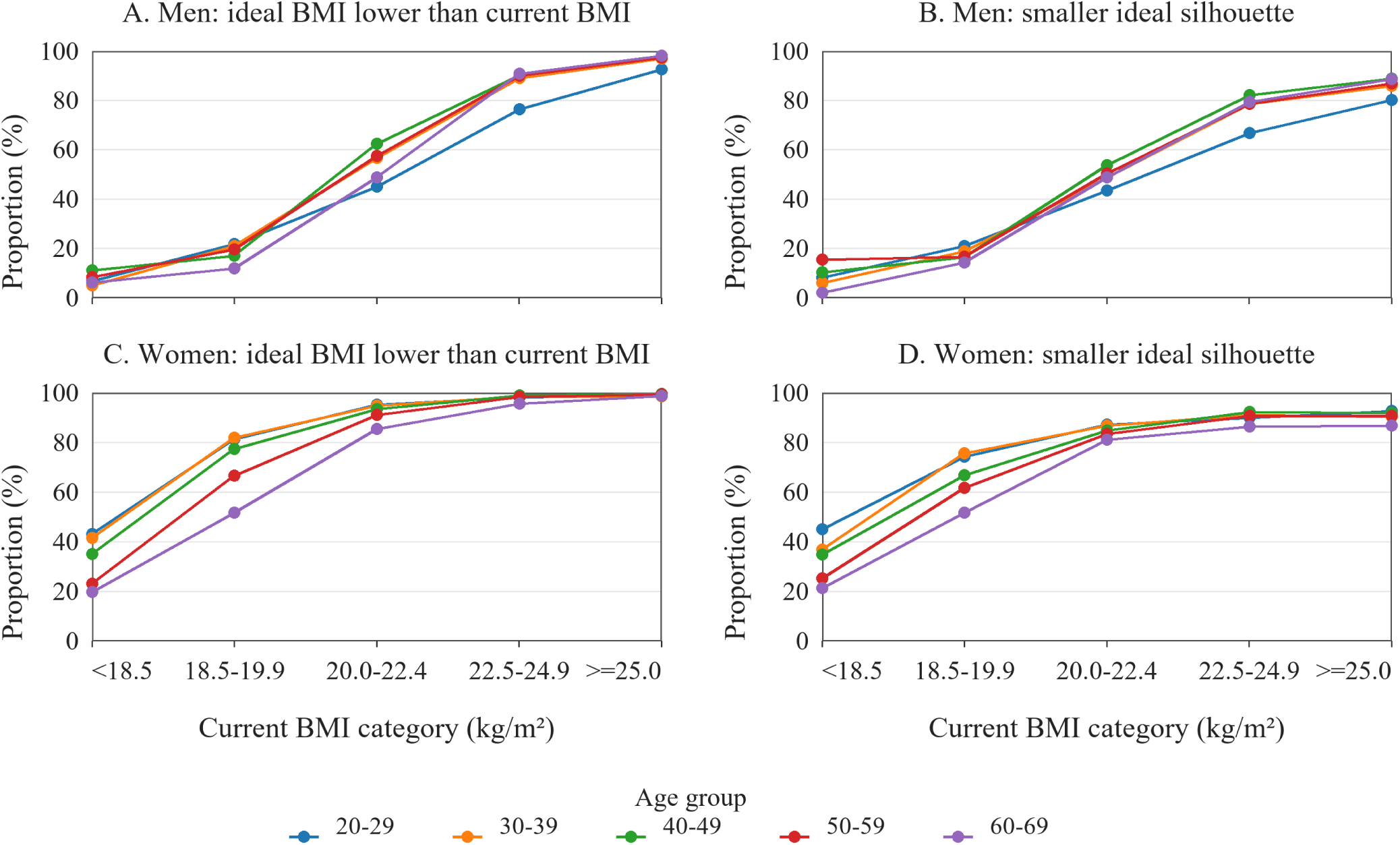
Relative thinness-oriented ideals by current BMI stratum, sex, and age group. Panels show the proportions of participants with lower-BMI ideals (A, C) or smaller ideal silhouettes (B, D) across current BMI strata in men (A, B) and women (C, D). Lines represent age groups. Adult analyses were restricted to participants aged 20-69 years. Current BMI was categorized as <18.5, 18.5-19.9, 20.0-22.4, 22.5-24.9, and ≥25.0 kg/m².

Visual body-size ideals were assessed using sex-specific 9-figure Stunkard Figure Rating Scales.^27^ Participants selected the silhouette that best represented their perceived current body size and the silhouette that best represented their ideal body size. Silhouette scores ranged from 1 to 9, with higher scores indicating larger body size. Visual body-size orientation was categorized by comparing the ideal silhouette with the perceived current silhouette: smaller ideal silhouette, same ideal silhouette, or larger ideal silhouette.

In this study, thinness-oriented ideals refer to a relative preference for a lower BMI (ideal BMI lower than current BMI) or a smaller silhouette (ideal silhouette smaller than the perceived current silhouette). This term does not necessarily indicate a desire to attain an underweight BMI or disordered eating.

Variables designed for broader perception-related analyses, including BMI-calibrated silhouette discrepancy, are listed in Supplementary Table S1 but were not analyzed in the present report.

### Other questionnaire domains

The broader questionnaire was designed to support subsequent analyses of body size, body image, health behaviours, and age- and sex-specific health outcomes. The present manuscript focused on visual and numerical body-size ideals because these were core measures available across the full survey sample. Additional domains, including sociocultural appearance attitudes, eating attitudes, dietary behaviours, physical activity, health literacy, and sex-specific health items, are listed in Supplementary Table S1 for transparency but were not treated as primary predictors or outcomes in the present initial descriptive report. These domains were reserved for future hypothesis-driven analyses to keep the present manuscript focused on the initial descriptive mapping of core body-size ideals.

### Statistical analysis

Participants were described in six age groups: 15-19, 20-29, 30-39, 40-49, 50-59, and 60-69 years. Continuous variables are presented as medians and interquartile ranges, and categorical variables as n (%). Missing values were not imputed. In Table 1, variable-specific non-missing denominators were used for BMI-related medians and proportions; the N column gives the total number of participants in each sex-age group. In the analytic file, current BMI was missing for 167 participants and ideal BMI for 252 participants. Adult BMI-stratified analyses were restricted to participants aged 20-69 years because BMI-for-age references are more appropriate through age 19 years.^28^

**Table 1.** Participant characteristics and body-size measures by sex and age group.

| Sex | Age group | N | Current BMI, median (IQR) | Current BMI category, n (%) |  |  | Ideal BMI, median (IQR) | Ideal BMI <18.5, n (%) | Current silhouette, median (IQR) | Ideal silhouette, median (IQR) |
| --- | --- | --- | --- | --- | --- | --- | --- | --- | --- | --- |
|  |  |  |  | <18.5 | 18.5-24.9 | ≥25.0 |  |  |  |  |
| Men | 15-19 | 730 | 20.5 (18.8-22.6) | 159 (21.9) | 489 (67.4) | 77 (10.6) | 20.6 (19.3-22.0) | 101 (14.0) | 4 (3-5) | 4 (3-5) |
|  | 20-29 | 1,594 | 21.4 (19.6-23.4) | 191 (12.1) | 1,186 (75.1) | 203 (12.8) | 21.2 (20.0-22.5) | 102 (6.5) | 4 (3-5) | 4 (3-5) |
|  | 30-39 | 1,700 | 22.4 (20.4-24.8) | 99 (5.8) | 1,190 (70.2) | 405 (23.9) | 21.5 (20.3-22.9) | 73 (4.3) | 5 (4-6) | 4 (3-5) |
|  | 40-49 | 2,142 | 23.0 (21.0-25.4) | 106 (5.0) | 1,427 (67.0) | 596 (28.0) | 21.6 (20.5-22.9) | 64 (3.0) | 5 (4-6) | 4 (3-5) |
|  | 50-59 | 2,137 | 23.3 (21.3-25.6) | 71 (3.3) | 1,408 (66.2) | 648 (30.5) | 22.0 (20.8-23.1) | 47 (2.2) | 5 (4-6) | 4 (3-5) |
|  | 60-69 | 1,791 | 23.4 (21.5-25.6) | 48 (2.7) | 1,203 (67.3) | 537 (30.0) | 22.0 (21.0-23.3) | 26 (1.5) | 5 (4-6) | 4 (4-5) |
| Women | 15-19 | 690 | 20.0 (18.4-21.6) | 186 (27.0) | 457 (66.3) | 46 (6.7) | 18.7 (17.7-20.0) | 324 (47.4) | 4 (4-5) | 3 (3-4) |
|  | 20-29 | 3,038 | 19.8 (18.5-21.5) | 750 (24.9) | 2,090 (69.3) | 178 (5.9) | 18.7 (17.8-19.8) | 1,324 (44.1) | 4 (3-5) | 3 (2-4) |
|  | 30-39 | 3,266 | 20.2 (18.8-22.4) | 680 (21.0) | 2,198 (67.8) | 364 (11.2) | 19.0 (18.0-20.2) | 1,216 (37.7) | 4 (4-5) | 3 (3-4) |
|  | 40-49 | 4,186 | 20.6 (18.8-22.9) | 831 (20.0) | 2,802 (67.4) | 522 (12.6) | 19.2 (18.2-20.4) | 1,324 (31.9) | 4 (4-5) | 3 (3-4) |
|  | 50-59 | 4,228 | 20.8 (18.9-23.1) | 820 (19.5) | 2,781 (66.2) | 602 (14.3) | 19.5 (18.5-20.8) | 1,088 (25.9) | 5 (4-6) | 3 (3-4) |
|  | 60-69 | 1,857 | 20.8 (18.9-23.1) | 373 (20.2) | 1,232 (66.9) | 237 (12.9) | 20.0 (18.8-21.2) | 367 (19.9) | 5 (4-5) | 4 (3-4) |

Values are median (IQR) or n (%). The N column gives the total number of participants in each sex-age group. BMI-related medians and proportions were calculated using variable-specific non-missing denominators; therefore, current BMI category counts may not sum to N when current BMI was missing, and denominators for ideal BMI <18.5 may differ where ideal BMI was missing. Current BMI categories were calculated from current BMI using cutoffs of <18.5, 18.5-24.9, and ≥25.0 kg/m<sup>2</sup>. Adult BMI cutoffs are shown descriptively for all age groups, including participants aged 15-19 years. Silhouette scores were treated descriptively as ordered numeric scores and are summarized as medians (IQRs). Percentages are unweighted. BMI, body mass index; IQR, interquartile range.

The main analysis first summarized BMI-status-specific numerical and visual orientations by sex and age group, using conventional adult BMI categories (underweight, normal weight, and obesity), with particular attention to normal-weight women, underweight women, and underweight men. To further characterize within-category gradients, we then examined the proportions of adults with relative thinness-oriented ideals (ideal BMI lower than current BMI; smaller ideal silhouette) and relative weight-gain-oriented ideals (ideal BMI higher than current BMI; larger ideal silhouette) by sex, age group, and finer current-BMI stratum. These proportions were intended to describe directional patterns rather than clinically meaningful thresholds of desired change. Among adults aged 20-69 years, 25,704 participants had valid values for both current and ideal BMI and were included in BMI-status-specific and finer-BMI-stratum analyses. For Table 2 and Figure 1 and 2, the row- or stratum-specific valid N was used as the denominator for all percentages. Analyses were conducted using IBM SPSS Statistics version 28 and Python.

**Table 2.**
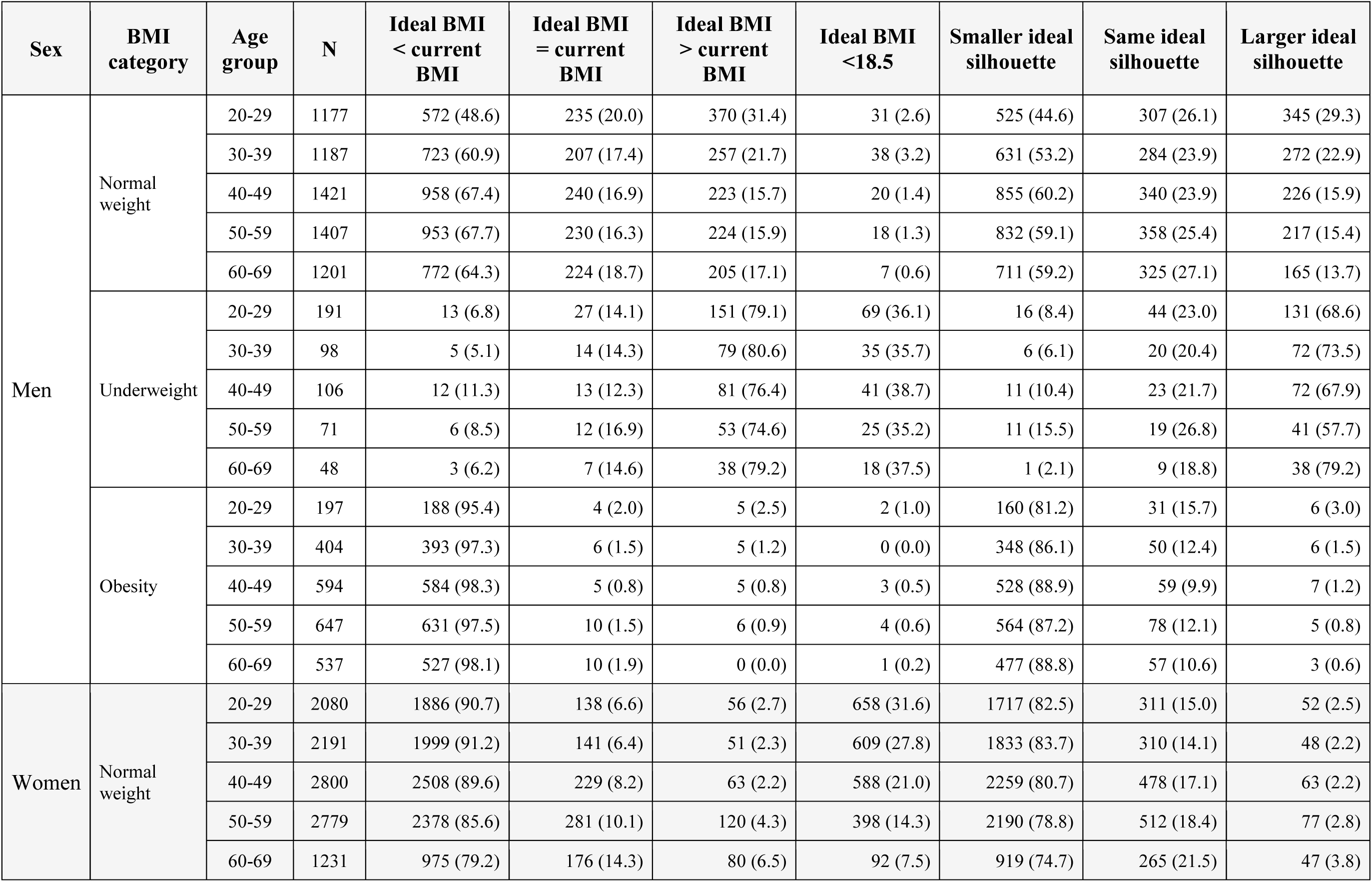
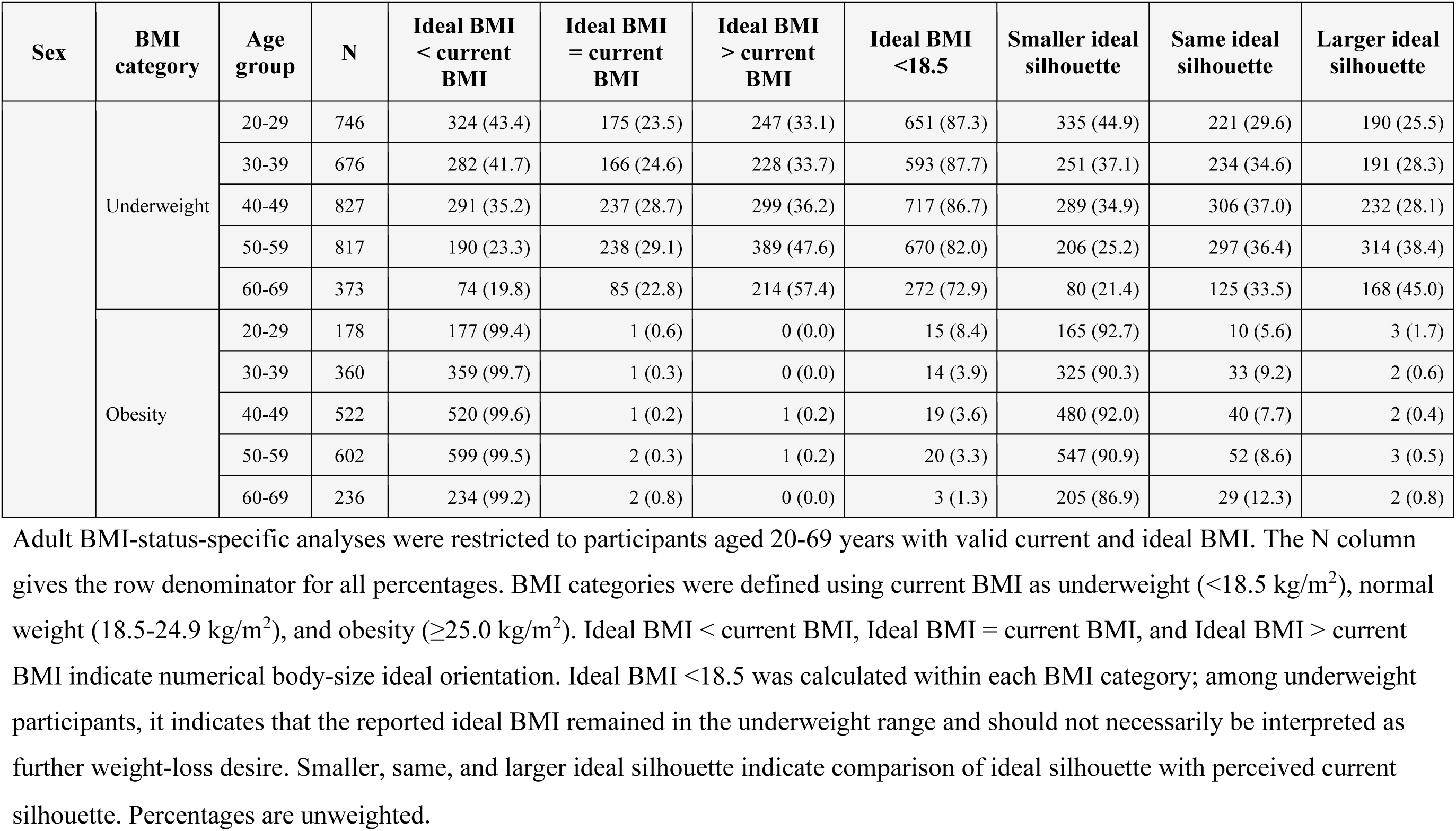
BMI-status-specific visual and numerical body-size ideals by sex and age group.

| Sex | BMI category | Age group | N | Ideal BMI < current BMI | Ideal BMI = current BMI | Ideal BMI > current BMI | Ideal BMI <18.5 | Smaller ideal silhouette | Same ideal silhouette | Larger ideal silhouette |
| --- | --- | --- | --- | --- | --- | --- | --- | --- | --- | --- |
| Men | Normal weight | 20-29 | 1177 | 572 (48.6) | 235 (20.0) | 370 (31.4) | 31 (2.6) | 525 (44.6) | 307 (26.1) | 345 (29.3) |
|  |  | 30-39 | 1187 | 723 (60.9) | 207 (17.4) | 257 (21.7) | 38 (3.2) | 631 (53.2) | 284 (23.9) | 272 (22.9) |
|  |  | 40-49 | 1421 | 958 (67.4) | 240 (16.9) | 223 (15.7) | 20 (1.4) | 855 (60.2) | 340 (23.9) | 226 (15.9) |
|  |  | 50-59 | 1407 | 953 (67.7) | 230 (16.3) | 224 (15.9) | 18 (1.3) | 832 (59.1) | 358 (25.4) | 217 (15.4) |
|  |  | 60-69 | 1201 | 772 (64.3) | 224 (18.7) | 205 (17.1) | 7 (0.6) | 711 (59.2) | 325 (27.1) | 165 (13.7) |
|  | Underweight | 20-29 | 191 | 13 (6.8) | 27 (14.1) | 151 (79.1) | 69 (36.1) | 16 (8.4) | 44 (23.0) | 131 (68.6) |
|  |  | 30-39 | 98 | 5 (5.1) | 14 (14.3) | 79 (80.6) | 35 (35.7) | 6 (6.1) | 20 (20.4) | 72 (73.5) |
|  |  | 40-49 | 106 | 12 (11.3) | 13 (12.3) | 81 (76.4) | 41 (38.7) | 11 (10.4) | 23 (21.7) | 72 (67.9) |
|  |  | 50-59 | 71 | 6 (8.5) | 12 (16.9) | 53 (74.6) | 25 (35.2) | 11 (15.5) | 19 (26.8) | 41 (57.7) |
|  |  | 60-69 | 48 | 3 (6.2) | 7 (14.6) | 38 (79.2) | 18 (37.5) | 1 (2.1) | 9 (18.8) | 38 (79.2) |
|  | Obesity | 20-29 | 197 | 188 (95.4) | 4 (2.0) | 5 (2.5) | 2 (1.0) | 160 (81.2) | 31 (15.7) | 6 (3.0) |
|  |  | 30-39 | 404 | 393 (97.3) | 6 (1.5) | 5 (1.2) | 0 (0.0) | 348 (86.1) | 50 (12.4) | 6 (1.5) |
|  |  | 40-49 | 594 | 584 (98.3) | 5 (0.8) | 5 (0.8) | 3 (0.5) | 528 (88.9) | 59 (9.9) | 7 (1.2) |
|  |  | 50-59 | 647 | 631 (97.5) | 10 (1.5) | 6 (0.9) | 4 (0.6) | 564 (87.2) | 78 (12.1) | 5 (0.8) |
|  |  | 60-69 | 537 | 527 (98.1) | 10 (1.9) | 0 (0.0) | 1 (0.2) | 477 (88.8) | 57 (10.6) | 3 (0.6) |
| Women | Normal weight | 20-29 | 2080 | 1886 (90.7) | 138 (6.6) | 56 (2.7) | 658 (31.6) | 1717 (82.5) | 311 (15.0) | 52 (2.5) |
|  |  | 30-39 | 2191 | 1999 (91.2) | 141 (6.4) | 51 (2.3) | 609 (27.8) | 1833 (83.7) | 310 (14.1) | 48 (2.2) |
|  |  | 40-49 | 2800 | 2508 (89.6) | 229 (8.2) | 63 (2.2) | 588 (21.0) | 2259 (80.7) | 478 (17.1) | 63 (2.2) |
|  |  | 50-59 | 2779 | 2378 (85.6) | 281 (10.1) | 120 (4.3) | 398 (14.3) | 2190 (78.8) | 512 (18.4) | 77 (2.8) |
|  |  | 60-69 | 1231 | 975 (79.2) | 176 (14.3) | 80 (6.5) | 92 (7.5) | 919 (74.7) | 265 (21.5) | 47 (3.8) |
|  | Underweight | 20-29 | 746 | 324 (43.4) | 175 (23.5) | 247 (33.1) | 651 (87.3) | 335 (44.9) | 221 (29.6) | 190 (25.5) |
|  |  | 30-39 | 676 | 282 (41.7) | 166 (24.6) | 228 (33.7) | 593 (87.7) | 251 (37.1) | 234 (34.6) | 191 (28.3) |
|  |  | 40-49 | 827 | 291 (35.2) | 237 (28.7) | 299 (36.2) | 717 (86.7) | 289 (34.9) | 306 (37.0) | 232 (28.1) |
|  |  | 50-59 | 817 | 190 (23.3) | 238 (29.1) | 389 (47.6) | 670 (82.0) | 206 (25.2) | 297 (36.4) | 314 (38.4) |
|  |  | 60-69 | 373 | 74 (19.8) | 85 (22.8) | 214 (57.4) | 272 (72.9) | 80 (21.4) | 125 (33.5) | 168 (45.0) |
|  | Obesity | 20-29 | 178 | 177 (99.4) | 1 (0.6) | 0 (0.0) | 15 (8.4) | 165 (92.7) | 10 (5.6) | 3 (1.7) |
|  |  | 30-39 | 360 | 359 (99.7) | 1 (0.3) | 0 (0.0) | 14 (3.9) | 325 (90.3) | 33 (9.2) | 2 (0.6) |
|  |  | 40-49 | 522 | 520 (99.6) | 1 (0.2) | 1 (0.2) | 19 (3.6) | 480 (92.0) | 40 (7.7) | 2 (0.4) |
|  |  | 50-59 | 602 | 599 (99.5) | 2 (0.3) | 1 (0.2) | 20 (3.3) | 547 (90.9) | 52 (8.6) | 3 (0.5) |
|  |  | 60-69 | 236 | 234 (99.2) | 2 (0.8) | 0 (0.0) | 3 (1.3) | 205 (86.9) | 29 (12.3) | 2 (0.8) |

Adult BMI-status-specific analyses were restricted to participants aged 20-69 years with valid current and ideal BMI. The N column gives the row denominator for all percentages. BMI categories were defined using current BMI as underweight ( $<18.5 \text{ kg/m}^2$ ), normal weight ( $18.5\text{-}24.9 \text{ kg/m}^2$ ), and obesity ( $\geq 25.0 \text{ kg/m}^2$ ). Ideal BMI < current BMI, Ideal BMI = current BMI, and Ideal BMI > current BMI indicate numerical body-size ideal orientation. Ideal BMI <18.5 was calculated within each BMI category; among underweight participants, it indicates that the reported ideal BMI remained in the underweight range and should not necessarily be interpreted as further weight-loss desire. Smaller, same, and larger ideal silhouette indicate comparison of ideal silhouette with perceived current silhouette. Percentages are unweighted.

**Figure 2.**
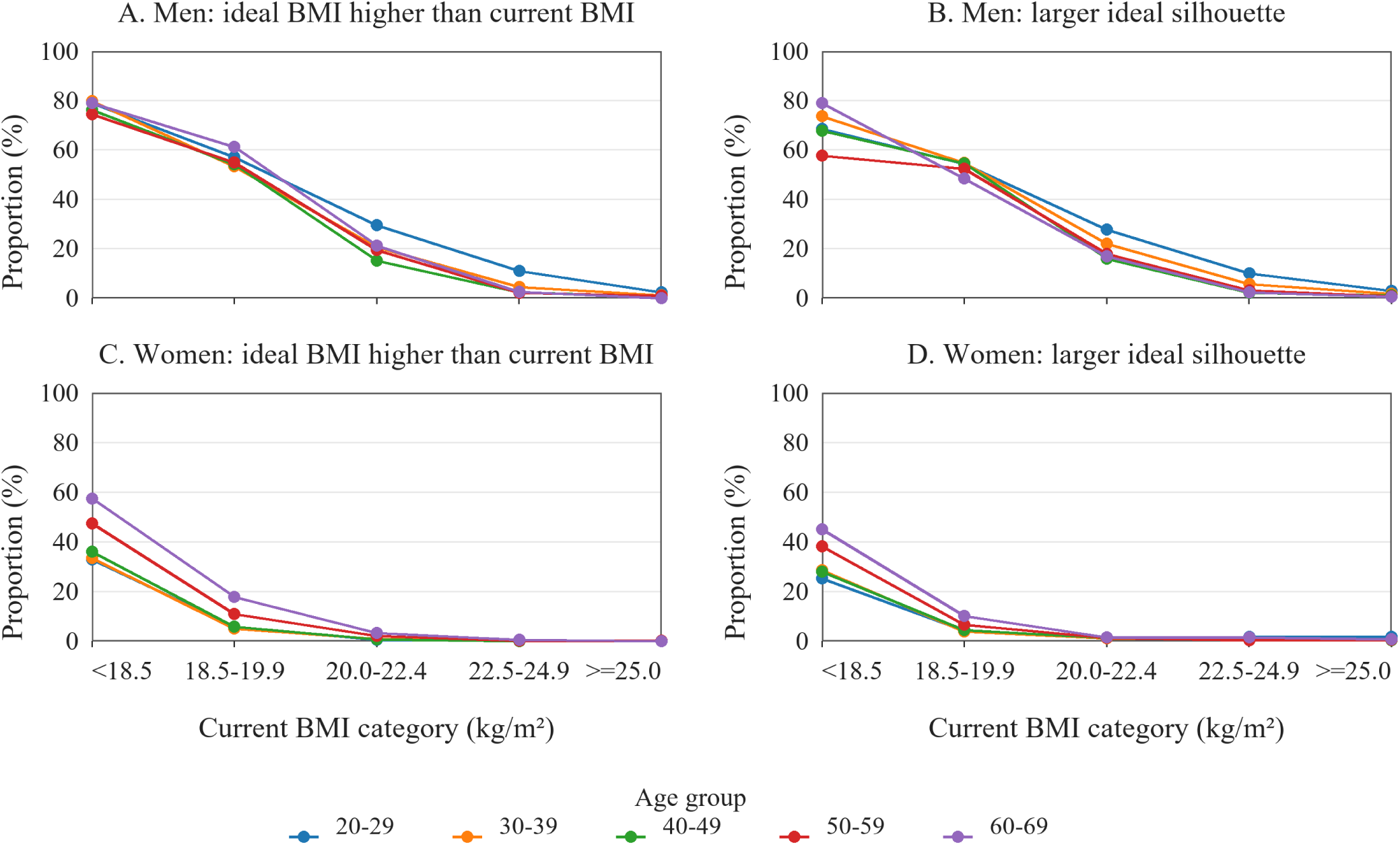
Relative weight-gain-oriented ideals by current BMI stratum, sex, and age group. Panels show the proportions of participants with higher-BMI ideals (A, C) or larger ideal silhouettes (B, D) across current BMI strata in men (A, B) and women (C, D). Lines represent age groups. Adult analyses were restricted to participants aged 20-69 years. Current BMI was categorized as <18.5, 18.5-19.9, 20.0-22.4, 22.5-24.9, and ≥25.0 kg/m².

### Ethics

The study was conducted in accordance with the Declaration of Helsinki and the Ethical Guidelines for Life Science and Medical Research Involving Human Subjects in Japan. The overall project was approved by the Research Ethics Committee of the Faculty of Medicine, Juntendo University (first approval no. E23-0305-M01; latest revised version no. E23-0305-M06). Electronic informed consent was obtained from all participants before participation. For participants younger than 18 years, assent and proxy consent from a parent or legal guardian were obtained in accordance with the study protocol.

## Results

### Participant characteristics

A total of 27,359 participants were included, comprising 10,094 men and 17,265 women distributed across six age groups from 15 to 69 years (Table 1). Median current BMI increased with age in both sexes, from 20.5 kg/m^2^ in men aged 15-19 years to 23.4 kg/m^2^ in men aged 60-69 years, and from 20.0 kg/m^2^ to 20.8 kg/m^2^ in women across the same age range. The proportion of participants with current BMI <18.5 kg/m^2^ was higher in women than in men within every adult age group, reaching 24.9% in women aged 20-29 years compared with 12.1% in men of the same age group. The proportion with current BMI ≥25.0 kg/m^2^ generally increased with age in both sexes and was higher in men than in women within each adult age group.

### Age- and sex-specific body-size measures

Age- and sex-specific medians of body-size ideals differed in direction between sexes (Table 1). In women, median ideal BMI was lower than median current BMI in every age group, including ages 15-19 years, and median ideal silhouette was smaller than median perceived current silhouette in every age group. In men, median ideal BMI was similar to or slightly lower than median current BMI depending on age, and median ideal silhouette was similar to or only slightly smaller than median perceived current silhouette. Ideal BMI <18.5 kg/m^2^ was reported by 47.4% of women aged 15-19 years and 44.1% of women aged 20-29 years, declining with age but still reported by 19.9% of women aged 60-69 years. In men, ideal BMI <18.5 kg/m^2^ was less common and declined more steeply with age, from 14.0% at ages 15-19 years to 1.5% at ages 60-69 years. Together, these age- and sex-specific medians indicate a more consistently thinness-oriented direction of body-size ideals in women than in men across the age range studied.

### BMI-status-specific ideals: normal-weight women, underweight women, and underweight men

BMI-status-specific estimates of numerical and visual ideals for adults are summarized in Table 2. Among normal-weight adult women, thinness-oriented ideals were common and persisted across all age groups: a smaller ideal silhouette was selected by 74.7% to 83.7%, and ideal BMI lower than current BMI was reported by 79.2% to 91.2%. The proportion reporting an absolute low-BMI ideal (ideal BMI <18.5 kg/m^2^) within this subgroup declined with age, from 31.6% at ages 20-29 years to 7.5% at ages 60-69 years, but remained non-negligible through midlife. Among underweight adult women, thinness-oriented indicators were also observed, particularly in younger age groups: a smaller ideal silhouette was selected by 44.9% at ages 20-29 years and decreased to 21.4% at ages 60-69 years, and ideal BMI lower than current BMI was reported by 43.4% and 19.8% in the same age groups, respectively. The proportion reporting ideal BMI <18.5 kg/m^2^ in this subgroup remained high across age groups (72.9%-87.7%), indicating that the reported ideal commonly remained within the underweight range itself. In contrast, underweight adult men frequently reported orientations in the opposite direction: ideal BMI higher than current BMI was reported by 74.6% to 80.6% across age groups, and a larger ideal silhouette was selected by 57.7% to 79.2%. Thus, underweight status was associated with thinness-oriented ideals in a meaningful subset of women, but with weight-gain-oriented ideals in most men.

### Patterns across finer current BMI strata

To further characterize within-category gradients, we next divided current BMI into five finer strata (<18.5, 18.5-19.9, 20.0-22.4, 22.5-24.9, and ≥25.0 kg/m^2^) and examined the distribution of thinness- and weight-gain-oriented ideals across these strata in adults aged 20-69 years, separately by sex and age group (Figures 1 and 2). In both sexes, the proportions reporting ideal BMI lower than current BMI and selecting a smaller ideal silhouette generally increased across current BMI strata (Figure 1), whereas the proportions reporting ideal BMI higher than current BMI and selecting a larger ideal silhouette decreased with higher current BMI (Figure 2). The shape of these distributions, however, differed by sex. In women, thinness-oriented ideals were already common within the low-normal (18.5-19.9 kg/m^2^) and mid-normal (20.0-22.4 kg/m^2^) BMI strata, and relative weight-gain-oriented ideals were largely confined to the underweight stratum but did not predominate even there. In men, thinness-oriented ideals were uncommon in the underweight and low-normal strata and rose mainly with higher current BMI, whereas relative weight-gain-oriented ideals were the most frequent orientation among underweight men. In the ≥25.0 kg/m^2^ stratum, lower-body-size-oriented responses were predominant in both sexes. Thus, fine-grained BMI stratification revealed a within-normal-range gradient in women that was not apparent from the three-category analysis alone.

## Discussion

In this large web-based survey of 27,359 Japanese adolescents and adults, we provided a high-resolution descriptive map of visual and numerical body-size ideals across sex, age, and current BMI strata in this Japanese web-panel sample. Three descriptive findings emerged. First, thinness-oriented body-size ideals were common among normal-weight adult women across all age groups and were already common within the low-normal (BMI 18.5-19.9 kg/m^2^) and mid-normal (20.0-22.4 kg/m^2^) BMI strata - a within-normal-range gradient that was not apparent from the conventional underweight/normal/obesity categorization alone. Second, the pattern in the underweight subgroup differed strikingly by sex: most underweight men preferred a larger or heavier body, whereas a subset of underweight women, particularly in younger age groups, preferred a smaller or lower one. Third, visual (silhouette-based) and numerical (BMI-based) indicators yielded broadly concordant group-level pictures of these patterns. The main contribution of this study is not the qualitative observation that Japanese women with normal or low BMI may desire a smaller body, which has been described in previous Japanese research; rather, it is the simultaneous, high-resolution mapping of these patterns by sex, age, and finer BMI stratum in a single large sample, which exposes a within-normal-range gradient with potential relevance to public-health priorities at both ends of Japanese adulthood.

The high frequency of thinness-oriented ideals among normal-weight Japanese women confirms and extends previous Japanese research. Prior studies have shown that Japanese adolescent girls and young adult women, including those who are normal weight or underweight, frequently report a desire for a smaller body.^16-18^ Our findings show that this orientation is not confined to young adulthood: an ideal BMI lower than current BMI was reported by 79·2–91·2% of normal-weight women across the adult age groups, including women aged 50–69 years. This finding is broadly consistent with international evidence that body dissatisfaction can persist into midlife and later adulthood.^5,7,20^ A more novel finding, enabled by finer BMI stratification, was that thinness-oriented ideals were already common among Japanese women in the low-normal and mid-normal BMI strata. This within-normal-range gradient was not apparent in the conventional three-category analysis and shows that such ideals are not confined to women with elevated current BMI. Notably, the prevalence of underweight (BMI <18·5 kg/m²) among Japanese women in their 20s rose markedly from approximately 15% in the 1980s to 20–25% by the 1990s and has remained elevated.^10,29^ The persistence of thinness-oriented ideals across the adult age range in the present study suggests that health education addressing body image may be relevant beyond young adulthood.

The opposite-direction pattern observed in underweight men — a preference for a larger ideal silhouette in 57·7-79·2% and a higher ideal BMI in 74·6-80·6% — is consistent with the broader literature on male body image, which has documented that low body size can be experienced as undesirable in men and that male body dissatisfaction often involves a wish for a more muscular and heavier body rather than a smaller one.^21,22^ However, the present silhouette- and BMI-based measures captured body-size direction only and could not distinguish a desire for greater muscularity or lean mass from a desire for greater body fat. Cross-national studies have similarly reported that men in modern industrialised societies tend to desire a body with substantially greater lean mass than they currently have or perceive themselves to have.^30^ Against this male pattern, our finding that a subset of underweight Japanese women still preferred a smaller body or a lower BMI, particularly in the youngest age group (44·9% of underweight women aged 20-29 years selected a smaller ideal silhouette), stands in sharper relief. Within underweight women, ideal BMI <18·5 kg/m2 was reported by 72·9–87·7% across age groups, indicating that the reported ideal commonly remained within the underweight range itself rather than reflecting an intention to gain weight into the normal range. Together, these contrasting patterns emphasize the need to consider sex and current BMI status jointly when interpreting body-size ideals.

A distinctive feature of our findings is the persistence of thinness-oriented ideals among normal-weight women well into midlife and early older adulthood. Even at ages 60-69 years, 74·7% of normal-weight women selected a smaller ideal silhouette and 79·2% reported an ideal BMI lower than their current BMI; in the underweight subgroup of the same age range, the reported ideal BMI remained within the underweight range in 72·9% of women. These findings are consistent with international evidence that body dissatisfaction does not dissolve with age in women,^5,7,20^ but they are particularly important in the Japanese context because low body weight and undernutrition in later adulthood are associated with frailty, sarcopenia, malnutrition, and increased all-cause mortality.^2,4,13,14^ The present cross-sectional data do not determine whether thinness-oriented ideals lead to later undernutrition or functional outcomes. However, they identify a descriptive pattern that may be relevant when designing future studies and public-health messages about adequate nutrition and muscle maintenance in midlife and early older adulthood.

These descriptive results may be relevant to current Japanese public-health priorities at both ends of adulthood. Among young women, Japan has the highest prevalence of underweight in the OECD,^9^ and approximately 20% of women in their 20s and 30s have a BMI <18·5 kg/m^2^;^10^ this clinical and population reality is being addressed through the recently proposed Female Underweight/Undernutrition Syndrome framework^11^ and as a population-level target within Health Japan 21 (third term).^15^ Our data do not establish that body-size ideals cause underweight, but they show that relative thinness-oriented ideals are already common at the low-normal end of the BMI distribution. Future longitudinal, behavioural, and intervention studies should test whether and how these ideals relate to weight-control behaviours, nutritional status, and movement into or out of underweight. At the other end of adulthood, Health Japan 21 (third term) also targets the reduction of low-nutrition tendency (BMI ≤20 kg/m2) among adults aged ≥65 years;^15^ our observation that thinness-oriented ideals persisted among normal-weight and underweight women aged 60-69 years suggests that body-image considerations may be one component to examine in strategies aimed at preventing frailty and sarcopenia in later life.

This study has several strengths. The analysis was based on 27,359 participants from all 47 Japanese prefectures, included both sexes across an age range spanning adolescence to early older adulthood, and assessed body-size ideals using both silhouette-based and BMI-based measures. The combined sample size enabled BMI-status-specific descriptions of subgroups - including underweight and obese adults of both sexes - that are typically underpowered in smaller surveys, and supported finer BMI stratification capable of detecting within-normal-range gradients. Several limitations should nonetheless be noted. First, the study was cross-sectional, so age-related patterns should not be interpreted as within-person developmental trajectories. Second, selection and non-response bias could not be quantified because participants were recruited from a commercial web panel and vendor-side recruitment metrics were unavailable.

Given the unweighted sample, the findings should not be interpreted as nationally representative prevalence estimates, and their generalisability beyond Japan may be limited. Third, height, current weight, and ideal weight were self-reported, which may have introduced measurement error in the derived BMI variables; the direction and magnitude of this error could not be quantified. Fourth, BMI-status-specific subgroup analyses were restricted to participants aged 20–69 years because BMI-for-age references are more appropriate for those aged 15–19 years.²⁸ Adult BMI cutoffs were used only for descriptive purposes for participants aged 15–19 years in Table 1. Fifth, the directional categories (for example, ideal BMI lower than current BMI) did not quantify the magnitude of desired BMI change, motivations for that change, or pathological thinness desire. In particular, among participants with BMI ≥25·0 kg/m², an ideal BMI lower than the current BMI may partly reflect health-motivated weight reduction and may not necessarily indicate a pathological desire for thinness. Sixth, the present initial report did not analyse the broader behavioural, psychological, nutritional, or sex-specific health domains listed in Supplementary Table S1, which were reserved for future hypothesis-driven analyses.

In conclusion, within this large demographically allocated, unweighted Japanese web-panel sample, visual and numerical thinness-oriented body-size ideals were common among adult women, persisted across all adult age groups, and were already common within the low-normal and mid-normal BMI strata - a within-normal-range gradient that emerged only when current BMI was examined at finer resolution. The pattern in the underweight subgroup diverged sharply by sex, with most underweight men preferring a larger or heavier body and a subset of underweight women, particularly those in their 20s, preferring a smaller or lower one. These findings provide a high-resolution descriptive map of body-size ideals across sex, age, and current BMI strata in a demographically allocated Japanese web-panel sample. Future studies should test whether these ideals are associated with weight-control behaviours, nutritional status, body composition, and health outcomes, and should examine whether body-image-sensitive health messages are useful for reducing female underweight and supporting adequate nutrition and healthy aging in midlife and early older adulthood.

## Data sharing statement

The deidentified participant-level data used in this study are not publicly available. Requests for data will be considered by the corresponding author, subject to approval by the relevant ethics committee and institution and the execution of an appropriate data-use agreement.

## Contributors

HT and YT conceptualised and designed the study and developed the methodology. HT planned and conducted the web-based survey and was responsible for project administration. HT, SS, KF, MK, and YA curated and managed the data. HT, KF, and YT performed the formal analyses. HT and YT interpreted the results, prepared the visualisations, and drafted the original manuscript. HT and YT supervised the study. YY and YT acquired funding and provided the resources required for the study. HT, SS, KF, MK, YA, and YT directly accessed the underlying data; HT, KF, and YT verified the underlying data and analytic results. All authors critically reviewed and edited the manuscript, approved the final version, and accepted responsibility for the decision to submit it for publication.

## Acknowledgments and role of the funding source

This work was supported by the Cabinet Office of Japan Strategic Innovation Promotion Program (SIP), "Inclusive Community Platform" (grant no. SIPCP23C02). The funder had no role in the study design; data collection, analysis, or interpretation; preparation of the manuscript; or the decision to submit the manuscript for publication.

## Declaration of use of AI and AI-assisted technologies in the writing process

During preparation of this manuscript, the authors used OpenAI Codex to assist with formatting the manuscript to journal requirements and with drafting clearly identified compliance text. The authors reviewed and edited the output and take full responsibility for the content of the publication.

## References

1 Guh DP, Zhang W, Bansback N, et al. The incidence of co-morbidities related to obesity and overweight: A systematic review and meta-analysis. BMC Public Health. 2009;9:88.

2 Bales CW, Ritchie CS. Sarcopenia, weight loss, and nutritional frailty in the elderly. Annu Rev Nutr. 2002;22:309–323.

3 Hunter GR, Plaisance EP, Fisher G. Weight loss and bone mineral density. Curr Opin Endocrinol Diabetes Obes. 2014;21(5):358–362.

4 Chen LK, Woo J, Assantachai P, Auyeung TW, Chou MY, Iijima K, et al. Asian Working Group for Sarcopenia: 2019 consensus update on sarcopenia diagnosis and treatment. J Am Med Dir Assoc. 2020;21(3):300–307.e2.

5 Tiggemann M, Lynch JE. Body image across the life span in adult women: the role of self-objectification. Dev Psychol. 2001;37(2):243–253.

6 Stice E, Marti CN, Durant S. Risk factors for onset of eating disorders: Evidence of multiple risk pathways from an 8-year prospective study. Behav Res Ther. 2011;49(10):622–627.

7 Quittkat HL, Hartmann AS, Dusing R, Buhlmann U, Vocks S. Body dissatisfaction, importance of appearance, and body appreciation in men and women over the lifespan. Front Psychiatry. 2019;10:864.

8 Bodega P, de Cos-Gandoy A, Fernández-Alvira JM, Fernández-Jiménez R. Body image and dietary habits in adolescents: a systematic review. Nutr Rev. 2024;82(1):104–127.

9 NCD Risk Factor Collaboration (NCD-RisC). Worldwide trends in underweight and obesity from 1990 to 2022: a pooled analysis of 3663 population-representative studies with 222 million children, adolescents, and adults. Lancet. 2024;403(10431):1027–1050.

10 Ministry of Health, Labour and Welfare. Summary of the results of the National Health and Nutrition Survey, Japan, 2023 [in Japanese]. Tokyo: Ministry of Health, Labour and Welfare; 2024. https://www.mhlw.go.jp/content/10900000/001338334.pdf. (Accessed May 11, 2026).

11 Tamura Y, Ogawa W, Ishii K, Ishigaki Y, Nagai N, Hirota Y, et al.; Working Group on Female Underweight/Undernutrition Syndrome; The Japan Society for the Study of Obesity. Female Underweight/Undernutrition Syndrome (FUS): an emerging health concept in premenopausal women-secondary publication (English translation of the Japanese statement). J Obstet Gynaecol Res. 2026;52:e70201. doi:10.1111/jog.70201.

12 Cabinet Office, Government of Japan. Annual Report on the Ageing Society [Summary] FY2025. Tokyo: Cabinet Office; 2025.

13 Fried LP, Tangen CM, Walston J, Newman AB, Hirsch C, Gottdiener J, et al. Frailty in older adults: evidence for a phenotype. J Gerontol A Biol Sci Med Sci. 2001;56(3):M146–M156.

14 Tamakoshi A, Yatsuya H, Lin Y, Tamakoshi K, Kondo T, Suzuki S, et al. BMI and all-cause mortality among Japanese older adults: findings from the Japan Collaborative Cohort Study. Obesity (Silver Spring). 2010;18(2):362–369. doi:10.1038/oby.2009.190.

15 Ministry of Health, Labour and Welfare. Overview of Health Japan 21 (the third term). Tokyo: Ministry of Health, Labour and Welfare; 2023. Available from: https://www.mhlw.go.jp/content/10904750/001158810.pdf. (Accessed May 11, 2026.)

16 Chisuwa N, O’Dea JA. Body image and eating disorders amongst Japanese adolescents: a review of the literature. Appetite. 2010;54(1):5–15.

17 Yasuda T. Desire for thinness among young Japanese women from the perspective of objective and subjective ideal body shape. Sci Rep. 2023;13:14129.

18 Suzuki T. Body image in female high school students: Reexamination with new figural stimuli. Jpn J Psychol. 2023;94(3):240–246. doi:10.4992/jjpsy.94.22309.

19 Frenzel A, Binder H, Walter N, et al. The aging human body shape. npj Aging Mech Dis. 2020;6:5.

20 Lewis DM, Cachelin FM. Body image, body dissatisfaction, and eating attitudes in midlife and elderly women. Eat Disord. 2001;9(1):29–39.

21 McCabe MP, Ricciardelli LA. Body image dissatisfaction among males across the lifespan: a review of past literature. J Psychosom Res. 2004;56(6):675–685.

22 Blond A. Impacts of exposure to images of ideal bodies on male body dissatisfaction: a review. Body Image. 2008;5(3):244–250.

23 von Elm E, Altman DG, Egger M, Pocock SJ, Gotzsche P, Vandenbroucke JP. The Strengthening the Reporting of Observational Studies in Epidemiology (STROBE) statement: guidelines for reporting observational studies. PLoS Med. 2007;4(10):e296.

24 Eysenbach G. Improving the quality of Web surveys: the Checklist for Reporting Results of Internet E-Surveys (CHERRIES). J Med Internet Res. 2004;6(3):e34.

25 Statistics Bureau of Japan. Current Population Estimates as of October 1, 2022. Tokyo: Statistics Bureau of Japan; 2023. https://www.stat.go.jp/english/data/jinsui/2022np/index.html. (Accessed May 11, 2026).

26 Examination Committee of Criteria for ‘Obesity Disease’ in Japan, Japan Society for the Study of Obesity. New criteria for ‘obesity disease’ in Japan. Circ J. 2002;66(11):987–992.

27 Stunkard AJ, Sørensen T, Schulsinger F. Use of the Danish Adoption Register for the study of obesity and thinness. In: Kety SS, Rowland LP, Sidman RL, Matthysse SW, eds. The Genetics of Neurological and Psychiatric Disorders. Raven Press; 1983:115–120.

28 de Onis M, Onyango AW, Borghi E, Siyam A, Nishida C, Siekmann J. Development of a WHO growth reference for school-aged children and adolescents. Bull World Health Organ. 2007;85(9):660–667.

29 Takimoto H, Yoshiike N, Kaneda F, Yoshita K. Thinness among young Japanese women. Am J Public Health. 2004;94(9):1592–1595. doi:10.2105/AJPH.94.9.1592.

30 Pope HG Jr, Gruber AJ, Mangweth B, Bureau B, deCol C, Jouvent R, Hudson JI. Body image perception among men in three countries. Am J Psychiatry. 2000;157(8):1297–1301. doi:10.1176/appi.ajp.157.8.1297.

