## Supplementary for "Sex-, age-, and BMI-specific patterns of visual and numerical body-size ideals in Japan: a large cross-sectional web-based survey"

**Contents**

Supplementary Table S1. Broader questionnaire domains 3

Supplementary Table S2. CHERRIES-related reporting items for the web-based survey 4

Supplementary Figure S1. Geographic distribution of participants across the 47 prefectures of Japan 5

References 6

### Supplementary Table S1. Broader questionnaire domains

| Domain | Examples of measurements/items | Role in present manuscript |
|---|---|---|
| Sociodemographic characteristics | Age, sex, prefecture of residence, household size, household composition, occupation, work/school attendance, household income, medical history, and health check-up history. | Used to describe the sample; prefecture, sex, and age were used for stratified sampling allocation. |
| Current and ideal body size | Self-reported height, current weight, and ideal weight; calculated current and ideal BMI; current–ideal BMI discrepancy; and current BMI categories. | Core measures. |
| Visual body-size ideals | Perceived current and ideal body silhouettes, selected from sex-specific 9-figure Stunkard Figure Rating Scales[1]; ideal silhouette categorized as smaller than, the same as, or larger than the perceived current silhouette. | Core measures. |
| BMI-calibrated silhouette discrepancy | Sex- and age-specific BMI norms for the standard 9-figure Stunkard Figure Rating Scale reported by Bulik et al.[2], together with a study-derived BMI-to-silhouette mapping, used to compare the perceived current silhouette with the silhouette category expected from the participant's current BMI. | Supplementary perception-related indicator. |
| Body-image and appearance-related items | Body-shape satisfaction and concerns; experiences of positive or negative comments about one's body shape; reasons for the desired body shape; age at first concern about body weight; and weight-control experiences. | Contextual descriptive measures; not included in the main analyses. |
| Standardized or scale-derived psychological measures | Sociocultural Attitudes Towards Appearance Questionnaire-3 Japanese Short Version (SATAQ-3 JS) subscale scores[3,4]; Body Appreciation Scale-2 (BAS-2)[5,6] and Drive for Thinness Scale scores[7], administered in applicable questionnaire branches; Eating Attitudes Test-26 (EAT-26)[8,9]; Japanese eHealth Literacy Scale (eHEALS)[10,11]; Ten-Item Personality Inventory–Japanese version (TIPI-J)[12,13]; Rosenberg Self-Esteem Scale (RSES)[14,15]; and Quick Inventory of Depressive Symptomatology–Self-Report Japanese version (QIDS-SR-J)[16,17]. | Contextual measures; not included in the main analyses. |
| Diet and nutrition-related items | Short self-administered food frequency questionnaire for the Japan Public Health Center–based Prospective Study for the Next Generation (short FFQ-NEXT), assessing habitual food and beverage intake and enabling estimation of energy and nutrient intake[18]; healthy-eating consciousness, calorie and nutrient awareness, dietary concerns, meal patterns (including meal skipping and late-night eating), eating practices, food and nutrition information sources, food-choice scenarios, fatty-meat-related gastrointestinal discomfort, and Vitamin D Deficiency Questionnaire for Japanese (VDDQ-J)-derived items[19]. | Reserved for subsequent dietary and symptom analyses. |
| Physical activity and sedentary behaviour | Global Physical Activity Questionnaire–Japanese version (GPAQ-J), assessing work-, transport-, and leisure-time physical activity and sedentary time[20–22]; current exercise and sports participation; and exercise habits across educational and adult life stages. | Reserved for subsequent physical-activity analyses. |
| General health and nonspecific symptoms | History, reasons for, and measurement methods of bone-density testing; skin and hair concerns; fatigue/malaise, migraine or chronic headache, eyestrain, sleep deprivation/insomnia, cold sensitivity, perceived tendency to lose or gain weight, and appetite changes in response to stress or fatigue. | Reserved for subsequent symptom-burden and general-health analyses. |
| Female reproductive health and related symptom items | Menstrual status and reproductive history; menstrual pain, irregularity, and premenstrual syndrome/premenstrual dysphoric disorder-related symptoms, including the 17-item PMDD-related scale developed by Miyaoka et al.[23]; Menstrual Distress Questionnaire (MDQ)-derived items adapted by Odagawa et al.[24,25]; and menopausal symptom items from the Simplified Menopausal Index (SMI)[26]. | Reserved for subsequent female-specific health and reproductive analyses. |
| Male-specific health items | Aging Males' Symptoms (AMS) scale-related items[27,28]; Sexual Health Inventory for Men (SHIM) items[29,30]; and Erection Hardness Score (EHS)-related items[31,32]. | Reserved for subsequent male health and sexual-function analyses. |

### Supplementary Table S2. CHERRIES-related reporting items for the web-based survey

| Reporting item | Information available in this report/analytic file | Notes |
| --- | --- | --- |
| Survey mode and administrator | Web-based component of the Survey of Body Shape and Health Awareness in Japanese People, administered by Rakuten Insight, Inc., to registered panel members. | Main-text Methods. |
| Data-collection period | March 11 to March 25, 2024. | Main-text Methods. |
| Recruitment and allocation | Stratified web-panel allocation based on the Population Estimates of Japan as of October 1, 2022; first-stage allocation of 20,000 respondents across 564 prefecture-by-sex-by-age strata, followed by an additional 7,359 women aged 20–59 years. | Percentages in the manuscript are unweighted. The additional female allocation improves precision for female-focused strata, but pooled percentages should not be interpreted as official national prevalence estimates. |
| Consent and eligibility | Electronic informed consent was obtained before participation. For participants younger than 18 years, assent and proxy consent were obtained in accordance with the protocol. | Protocol-defined exclusion criteria are described in the main text. |
| Duplicate prevention | Duplicate submissions from the same individual were prevented by vendor-assigned unique monitor IDs. | No participant identifiers were included in the analytic file. |
| Completed records | The deidentified analytic file contained completed response records only. | Incomplete or page-by-page response information was not included in the analytic file. |
| Incentive | Web survey respondents received Rakuten points according to the survey vendor rules. | The amount and administration were managed by the vendor. |
| Unavailable vendor-side metrics | Invitation counts, view rates, participation rates, and page-by-page completion information were unavailable in the analytic file. | Response processes and selection bias therefore could not be fully assessed. |
| Reporting framework | The manuscript was prepared in accordance with STROBE[33] and with reference to CHERRIES recommendations[34] where applicable. | This table is intended to clarify available and unavailable web-survey reporting information. |

**Supplementary Figure S1. Geographic distribution of participants across the 47 prefectures of Japan**

| No. | Prefecture | Total | Men | Women | No. | Prefecture | Total | Men | Women |
|---|---|---|---|---|---|---|---|---|---|
| 1 | Hokkaido | 1068 | 388 | 680 | 25 | Shiga | 322 | 118 | 204 |
| 2 | Aomori | 264 | 98 | 166 | 26 | Kyoto | 564 | 201 | 363 |
| 3 | Iwate | 258 | 98 | 160 | 27 | Osaka | 1915 | 681 | 1234 |
| 4 | Miyagi | 512 | 189 | 323 | 28 | Hyogo | 1150 | 410 | 740 |
| 5 | Akita | 198 | 76 | 122 | 29 | Nara | 286 | 102 | 184 |
| 6 | Yamagata | 226 | 88 | 138 | 30 | Wakayama | 206 | 72 | 134 |
| 7 | Fukushima | 383 | 148 | 235 | 31 | Tottori | 130 | 48 | 82 |
| 8 | Ibaraki | 616 | 235 | 381 | 32 | Shimane | 144 | 56 | 88 |
| 9 | Tochigi | 414 | 161 | 253 | 33 | Okayama | 398 | 145 | 253 |
| 10 | Gunma | 414 | 155 | 259 | 34 | Hiroshima | 592 | 219 | 373 |
| 11 | Saitama | 1594 | 599 | 995 | 35 | Yamaguchi | 282 | 104 | 178 |
| 12 | Chiba | 1360 | 510 | 850 | 36 | Tokushima | 160 | 58 | 102 |
| 13 | Tokyo | 3285 | 1192 | 2093 | 37 | Kagawa | 208 | 78 | 130 |
| 14 | Kanagawa | 2044 | 763 | 1281 | 38 | Ehime | 283 | 104 | 179 |
| 15 | Niigata | 450 | 167 | 283 | 39 | Kochi | 148 | 56 | 92 |
| 16 | Toyama | 226 | 84 | 142 | 40 | Fukuoka | 1103 | 391 | 712 |
| 17 | Ishikawa | 256 | 94 | 162 | 41 | Saga | 182 | 66 | 116 |
| 18 | Fukui | 172 | 66 | 106 | 42 | Nagasaki | 272 | 101 | 171 |
| 19 | Yamanashi | 186 | 70 | 116 | 43 | Kumamoto | 358 | 131 | 227 |
| 20 | Nagano | 419 | 158 | 261 | 44 | Oita | 238 | 88 | 150 |
| 21 | Gifu | 414 | 153 | 261 | 45 | Miyazaki | 232 | 86 | 146 |
| 22 | Shizuoka | 756 | 286 | 470 | 46 | Kagoshima | 330 | 119 | 211 |
| 23 | Aichi | 1630 | 617 | 1013 | 47 | Okinawa | 332 | 123 | 209 |
| 24 | Mie | 379 | 142 | 237 | Total | | 27359 | 10094 | 17265 |

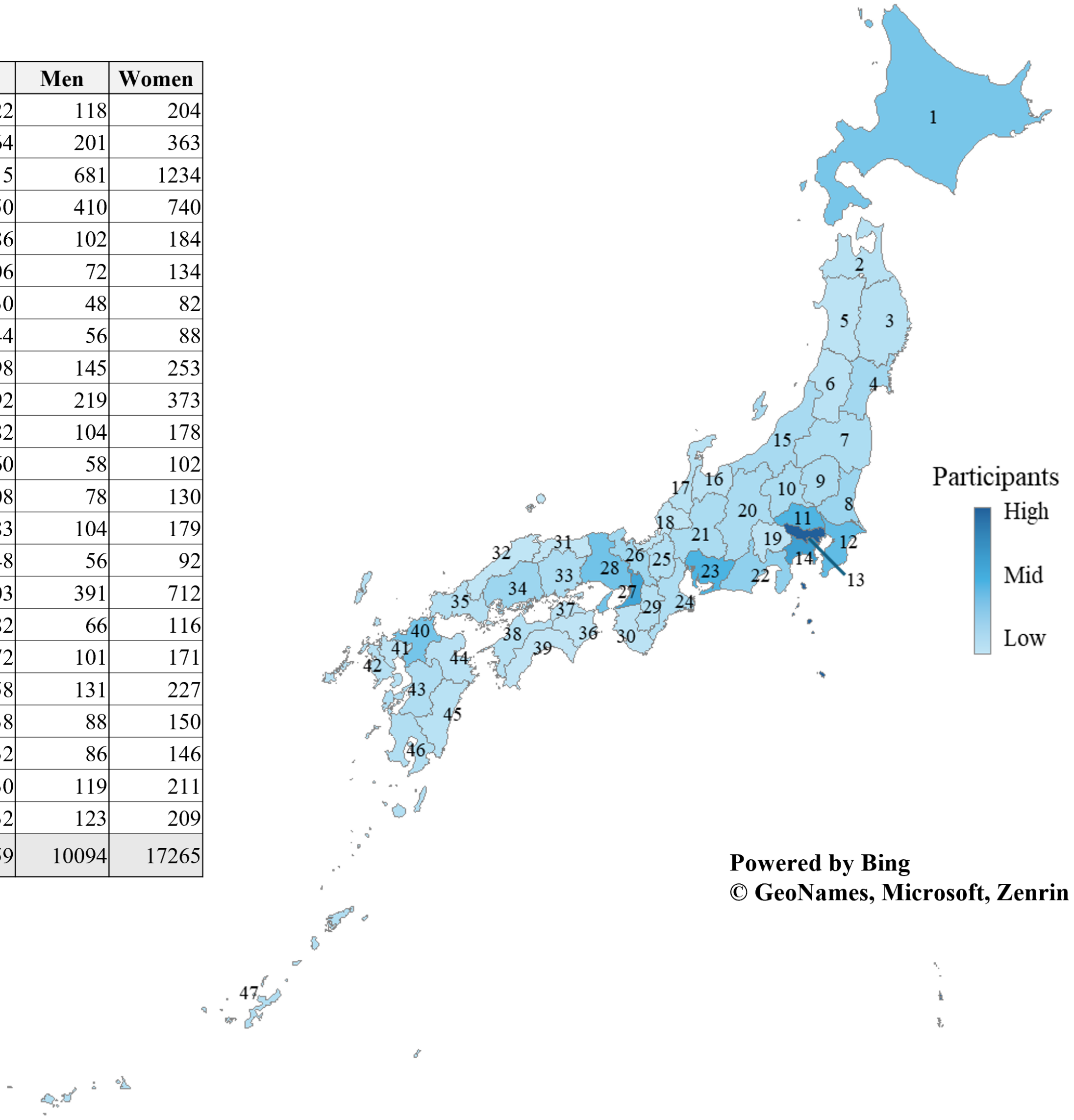


Prefecture numbers on the map correspond to those listed in the table. The table shows the total number of participants and the numbers of men and women in each prefecture. The map color intensity represents the relative number of participants in each prefecture. Map source: Bing Maps. © GeoNames, Microsoft, Zenrin.

### References


1 Stunkard AJ, Sorensen T, Schulsinger F. Use of the Danish Adoption Register for the Study of Obesity and Thinness. In: Kety SS, Rowland LP, Sidman RL, Matthysse SW, eds. The Genetics of Neurological and Psychiatric Disorders. Raven Press; 1983:115–120.

2 Bulik CM, Wade TD, Heath AC, Martin NG, Stunkard AJ, Eaves LJ. Relating body mass index to figural stimuli: population-based normative data for Caucasians. Int J Obes Relat Metab Disord. 2001;25(10):1517-1524.

3 Thompson JK, van den Berg P, Roehrig M, Guarda AS, Heinberg LJ. The Sociocultural Attitudes Towards Appearance Scale-3 (SATAQ-3): development and validation. Int J Eat Disord. 2004;35(3):293-304.

4 Yamamiya Y, Shimai S. Development of the Sociocultural Attitudes Towards Appearance Questionnaire-3 Japanese Short Version (SATAQ-3 JS) and establishment of its reliability and validity. Japanese Journal of Psychosomatic Medicine. 2012;52(1):54-63. [In Japanese]

5 Tylka TL, Wood-Barcalow NL. The Body Appreciation Scale-2: item refinement and psychometric evaluation. Body Image. 2015;12:53-67.

6 Namatame H, Uno K, Sawamiya Y. Development of Japanese version of the Body Appreciation Scale-2. Japanese Journal of Psychology. 2017;88(4):358-365. [In Japanese]

7 Matsumoto S, Kumano H, Sakano Y. Investigation of social influence on eating disorder tendencies and dieting behavior among female students. Behavior Therapy Research. 1999;25(2):11–23. doi:10.24468/jjbt.25.2_11. [In Japanese]

8 Garner DM, Olmsted MP, Bohr Y, Garfinkel PE. The Eating Attitudes Test: psychometric features and clinical correlates. Psychol Med. 1982;12(4):871-878.

9 Mukai T, Crago M, Shisslak CM. Eating attitudes and weight preoccupation among female high school students in Japan. J Child Psychol Psychiatry. 1994;35(4):677-688.

10 Norman CD, Skinner HA. eHEALS: The eHealth Literacy Scale. J Med Internet Res. 2006;8(4):e27. doi:10.2196/jmir.8.4.e27.

11 Mitsutake S, Shibata A, Ishii K, Okazaki K, Oka K. Developing the Japanese version of the eHealth Literacy Scale (eHEALS). Nihon Koshu Eisei Zasshi. 2011;58(5):361–371. doi:10.11236/jph.58.5_361. [In Japanese]

12 Gosling SD, Rentfrow PJ, Swann WB Jr. A very brief measure of the Big-Five personality domains. Journal of Research in Personality. 2003;37(6):504–528. doi:10.1016/S0092-6566(03)00046-1.

13 Oshio A, Abe S, Cutrone P. Development, reliability, and validity of the Japanese version of Ten Item Personality Inventory (TIPI-J). The Japanese Journal of Personality. 2012;21(1):40–52. doi:10.2132/personality.21.40.

14 Rosenberg M. Society and the Adolescent Self-Image. Princeton University Press; 1965.

15 Yamamoto M, Matsui Y, Yamanari Y. The structure of perceived aspects of self. Japanese Journal of Educational Psychology. 1982;30(1):64–68. doi:10.5926/jjep1953.30.1_64.

16 Rush AJ, Trivedi MH, Ibrahim HM, et al. The 16-item Quick Inventory of Depressive Symptomatology (QIDS), clinician rating (QIDS-C), and self-report (QIDS-SR): a psychometric evaluation in patients with chronic major depression. Biological Psychiatry. 2003;54(5):573–583. doi:10.1016/S0006-3223(02)01866-8.

17 Fujisawa D, Nakagawa A, Tajima M, et al. Cross-cultural adaptation of the Quick Inventory of Depressive Symptomatology–Self Report (QIDS-SR-J). Stress Science. 2010;25(1):43–52. [In Japanese]

18 Yokoyama Y, Takachi R, Ishihara J, et al. Validity of short and long self-administered food frequency questionnaires in ranking dietary intake in middle-aged and elderly Japanese in the Japan Public Health Center-Based Prospective Study for the Next Generation (JPHC-NEXT) Protocol Area. Journal of Epidemiology. 2016;26(8):420–432. doi:10.2188/jea.JE20150064.

19 Kuwabara A, Tsugawa N, Mizuno K, Ogasawara H, Watanabe Y, Tanaka K. A simple questionnaire for the prediction of vitamin D deficiency in Japanese adults (Vitamin D Deficiency Questionnaire for Japanese: VDDQ-J). J Bone Miner Metab. 2019;37:854-863.

20 World Health Organization. Global Physical Activity Questionnaire (GPAQ) analysis guide. Geneva: World Health Organization; 2021.

21 Japan Physical Activity Research Platform. Global Physical Activity Questionnaire, version 2: Japanese version (GPAQ ver. 2) [In Japanese]. Available from: https://paplatform.umin.jp/doc/gpaq.pdf. Accessed August 1, 2026.

22 Tanaka C, Reilly JJ, Tanaka S. Correlations between the Japanese version of the Global Physical Activity Questionnaire and accelerometer-measured physical activity and sedentary time in adult women and men. Journal of Activity, Sedentary and Sleep Behaviors. 2024;3:29. doi:10.1186/s44167-024-00069-1.

23 Miyaoka Y, Akimoto Y, Ueda K, Kamo T. The reliability and validity of the newly developed PMDD scale. Journal of Japanese Society of Psychosomatic Obstetrics and Gynecology. 2009;14(2):194-201. [In Japanese]

24 Odagawa H, Shirato N, Nagatsuka M, Chiba H, Kimura T, Okai T. An investigation of menstruation symptoms in young women by measuring modified MDQ scores. J Showa Med Assoc. 2008;68(3):155–161. doi:10.14930/jsma1939.68.155. [In Japanese]

25 Moos RH. The development of a menstrual distress questionnaire. Psychosom Med. 1968;30(6):853–867. doi:10.1097/00006842-196811000-00006.

26 Koyama T, Aso T. Kōnenki fujin ni okeru kanpō ryōhō: kanryakuka shita kōnenki shisū ni yoru hyōka [Kampo therapy for menopausal women: evaluation using a simplified menopausal index]. Sanfujinka Kampo Kenkyu no Ayumi. 1992;9:30–34. [In Japanese]

27 Heinemann LAJ, Zimmermann T, Vermeulen A, Thiel C, Hummel W. A new 'Aging Males' Symptoms' rating scale. Aging Male. 1999;2(2):105-114.

28 Kobayashi K, Hashimoto K, Kato R, et al. The aging males' symptoms scale for Japanese men: reliability and applicability of the Japanese version. Int J Impot Res. 2008;20:544-548.

29 Rosen RC, Cappelleri JC, Smith MD, Lipsky J, Peña BM. Development and evaluation of an abridged, 5-item version of the International Index of Erectile Function (IIEF-5) as a diagnostic tool for erectile dysfunction. International Journal of Impotence Research. 1999;11:319–326.

30 Kimoto Y, Ikeda S, Nagao K, et al. The New Japanese versions of International Index of Erectile Function (IIEF) and IIEF5. Jpn J Impot Res. 2009;24(3):295–308. [In Japanese]

31 Mulhall JP, Goldstein I, Bushmakin AG, Cappelleri JC, Hvidsten K. Validation of the Erection Hardness Score. J Sex Med. 2007;4(6):1626–1634. doi:10.1111/j.1743-6109.2007.00600.x.

32 Kimura M, Shimura S, Tai T, et al. A web-based survey of Erection Hardness Score and its relationship to aging, sexual behavior, confidence, and risk factors in Japan. Sex Med. 2013;1(2):76–86. doi:10.1002/sm2.15.

33 von Elm E, Altman DG, Egger M, Pocock SJ, Gotzsche PC, Vandenbroucke JP. The Strengthening the Reporting of Observational Studies in Epidemiology (STROBE) statement: guidelines for reporting observational studies. PLoS Med. 2007;4(10):e296.

34 Eysenbach G. Improving the quality of Web surveys: the Checklist for Reporting Results of Internet E-Surveys (CHERRIES). J Med Internet Res. 2004;6(3):e34.
